# Staffable ICU early-warning with calibrated multi-centre validation and alert-budget guardrails

**DOI:** 10.1101/2025.10.30.25339143

**Authors:** Sanwal Ahmad Zafar, Wei Qin, Toufique Ahmed Soomro, Areeba Ali Khan, Muhammad Salman Faisal

## Abstract

We present a staffable ICU early-warning system with calibrated multi-centre validation and alert-budget guardrails. A multi-horizon mortality model was developed on a large public ICU database and externally validated across independent hospitals at fixed post-admission landmarks (6/12/18/24 h). Evaluation was prespecified to emphasize actionability: discrimination, calibration (slope/intercept, Brier, ECE), decision-curve net benefit, and operating-point performance. To make deployment resource-aware, we translate threshold choice into **alerts per 100 admissions** and map alerts to **clinician review time** and **net monetary benefit** (NMB), yielding guardrails that bound workload while preserving utility. We further provide a **silent-trial** readout that estimates lead time prior to activation, supporting governance and cautious piloting. Across centres and horizons, the model maintained strong discrimination, tight calibration, and positive net benefit at clinically relevant thresholds. Guardrail selection produced **workload-constrained** operating points with stable subgroup performance. By coupling calibrated multicentre validation with alert-budget guardrails and deployment-proximal readouts, this study reframes ICU early-warning from maximizing AUC to delivering a **staffable, value-aware** program ready for monitored roll-out and periodic recalibration.

## Introduction

Clinical deterioration remains a major driver of morbidity, mortality, and resource use in the ICU. Machine-learning (ML) early-warning systems (EWS) have shown promising discrimination in retrospective studies, yet bedside usefulness depends as much on calibrated risk and actionable operating points as on rank order performance. Contemporary reporting guidance emphasizes complementing AUROC with calibration, clinical decision utility, and transparent description of intended use and thresholding, together with clear assessments of bias and applicability for ML-based prediction [1, 2]. In parallel, health-economic guidance encourages aligning evaluation with the decision context and explicitly considering opportunity costs when models trigger human review or downstream actions [3–5].

Two gaps limit translation. First, most EWS reports do not connect operating thresholds to staffable alert programs or quantify the workload those thresholds imply. Without an alert-budget perspective, even well-calibrated models can overwhelm teams with low-value alerts or underperform if set too conservatively. Second, deployment-proximal evaluation is often missing: before activation, services benefit from “silent-trial” assessment to estimate alert volume, timeliness, and missed events under local case-mix and staffing, and to confirm that thresholds deliver the intended balance of sensitivity, precision, and net clinical value.

We externally validate a pragmatic ICU mortality model at fixed landmarks (6, 12, 18, and 24 h), following current reporting and risk-of-bias recommendations [1, 2]. We report discrimination, calibration, decision-curve utility, and operating points carried unchanged to external data. We also introduce an alert-budget guardrail mapped to staff time, a concise NMB readout following economic reporting guidance [3, 4], and examine temporal stability and site-level synthesis. Additionally, we include SHAP summaries for transparency **(Figure 1)**.

**Figure 1.**
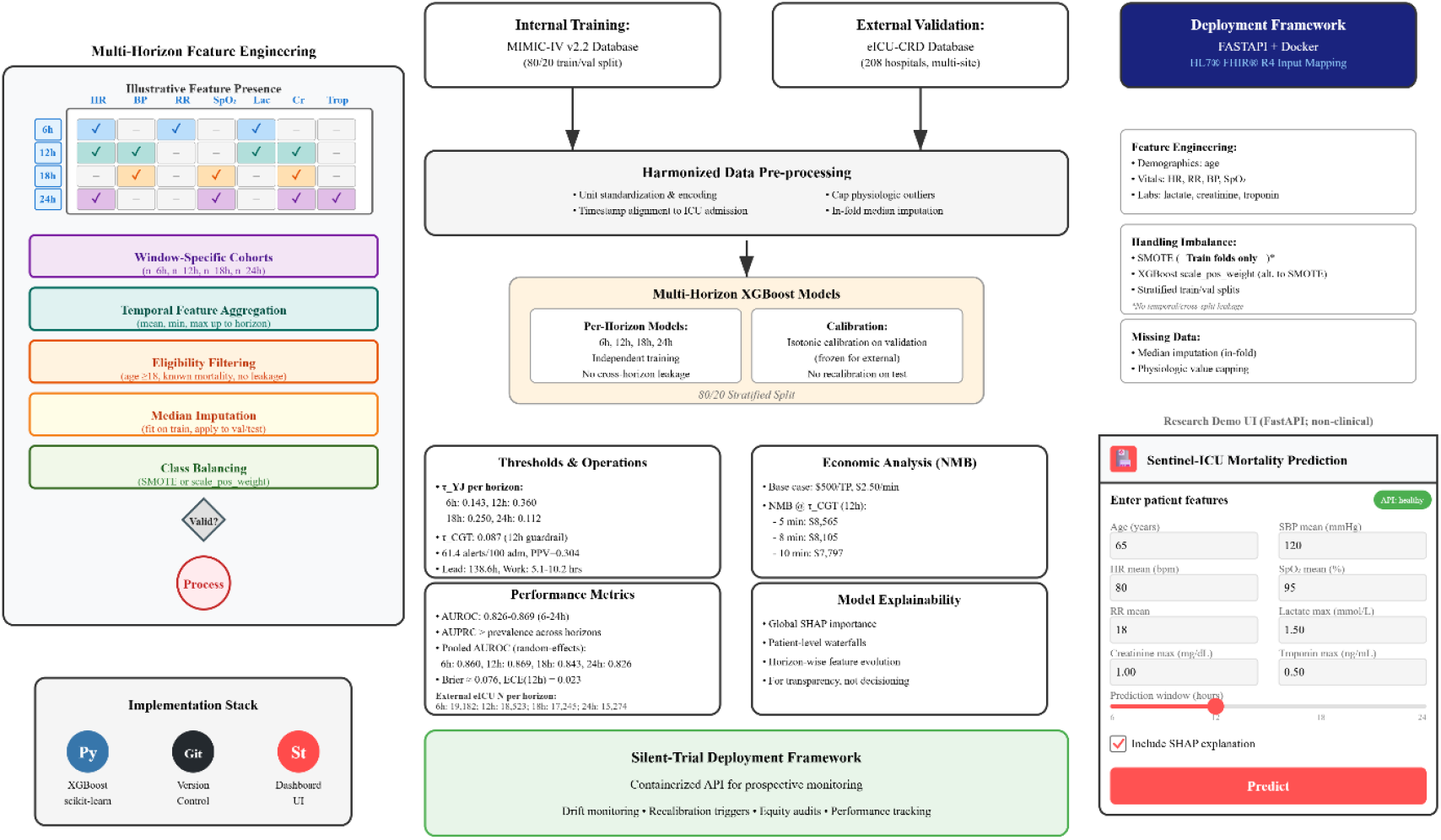
Sentinel-ICU pipeline overview. Model development workflow showing feature engineering at four horizons (6/12/18/24h), XGBoost modeling with isotonic calibration, dual thresholding strategy (τ_γj_ for accuracy, τ_*CGT*_for operational planning), and deployment framework. Key 12h metrics: Brier=0.076, ECE=0.023.

Our prespecified outcomes are: **(i)** discrimination and calibration at each landmark; **(ii)** decision-curve net benefit and horizon-specific operating-point performance; **(iii)** an operational readout at the 12-h guardrail linking alert rates to review workload and NMB; **(iv)** temporal stability and site-level heterogeneity; and **(v)** subgroup summaries to support equity and governance considerations. Taken together, the study is designed to move beyond “high AUROC” toward staffable, budget-constrained, and auditable ICU decision support, with all assumptions and threshold selection fully explained in Methods and outcomes reported concisely in result section.

## MATERIALS AND METHODS

### Operating thresholds and workload mapping

We used two thresholds. τ_γj_ (Youden’s J on calibrated risks, fixed from internal validation) for reporting accuracy metrics, and τ_*CGT*_(clinical guardrail threshold) set to meet a target alert budget (alerts per 100 admissions). Figure 1 (Thresholds & Operations) illustrates the roles of τ_γj_ and τ_CGT_We report accuracy at τ_γj_ and operational workload/economics at τ_*CGT*_, without model refitting or horizon leakage. Both thresholds were prespecified on the MIMIC-IV internal validation split and then carried unchanged to eICU-CRD; no threshold tuning was performed on external data. Alerts/100 were converted to minutes and clinician-hours/100 using 5-10 min/alert (Table 1).

**Table 1.**
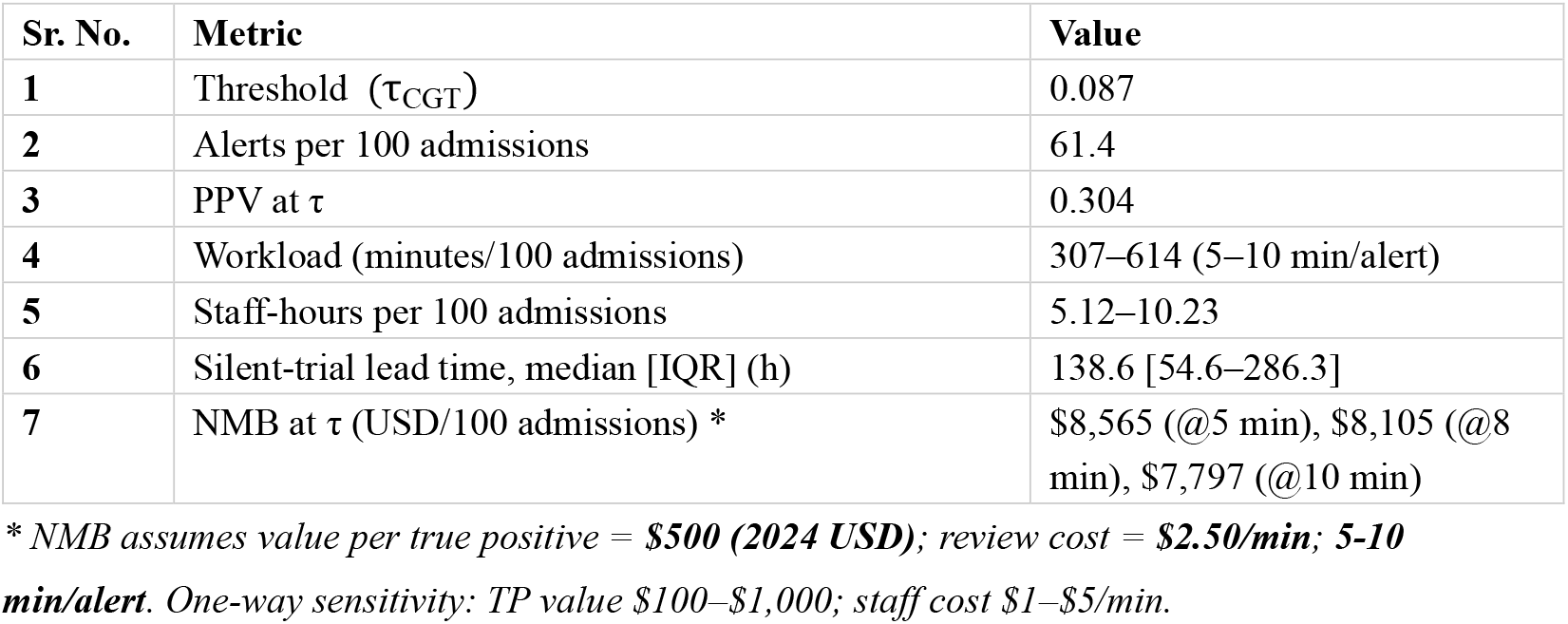
Operational Summary at 12 h (τ_CGT_)

| Sr. No. | Metric | Value |
| --- | --- | --- |
| 1 | Threshold ($\tau_{CGT}$) | 0.087 |
| 2 | Alerts per 100 admissions | 61.4 |
| 3 | PPV at $\tau$ | 0.304 |
| 4 | Workload (minutes/100 admissions) | 307–614 (5–10 min/alert) |
| 5 | Staff-hours per 100 admissions | 5.12–10.23 |
| 6 | Silent-trial lead time, median [IQR] (h) | 138.6 [54.6–286.3] |
| 7 | NMB at $\tau$ (USD/100 admissions) * | \$8,565 (@5 min), \$8,105 (@8 min), \$7,797 (@10 min) |

\* NMB assumes value per true positive = **\$500 (2024 USD)**; review cost = **\$2.50/min; 5-10 min/alert**. One-way sensitivity: TP value \$100–\$1,000; staff cost \$1–\$5/min.

### Clinical utility, economic evaluation and sub-group performance

Clinical utility was summarized by decision-curve net benefit (thresholds 0.01-0.30; treat-all/none comparators) and by a simple net monetary benefit (NMB) scenario (Figure 3C). Alert counts were converted to reviewer workload as alerts × minutes per alert (5-10) (Table 1), expressed as clinician-hours per 100 admissions, following CHEERS 2022 reporting guidance [3–5]. This approach follows established workload conversion and staffing equivalence guidelines [4, 6]. NMB reported with base-case assumptions; full formula in paragraph S11. At the prespecified 12-h threshold we report sensitivity and PPV (95% CIs) across sex, age, and admission source with the threshold fixed; full tables/definitions are in S12-S13 and Table S5.

### Software and Reproducible Tools

To reduce leakage and overfitting, we froze the feature set and hyperparameters before external testing. We also provide two runnable tools: Supplementary Tool S1, a lightweight FastAPI mortality-inference service and Supplementary Tool S2, an NMB forecaster that maps operating points. Frozen features, hash-locked splits, and repo tools, as well as supplementary tools and endpoints documented in S14.

### Study Cohorts and External Validation

External validation in eICU-CRD used four post-admission windows (6/12/18/24 h) with window-specific inclusion; thresholds, workload conversion, decision-curve/NMB, and silent-trial definitions follow Methods, details in paragraphs S2-S4. We computed discrimination (AUROC/AUPRC), calibration (reliability curves, ECE/Brier, slope/CITL (calibration-in-the-large)), decision-curve net benefit [6], NMB, and subgroup performance (sex, age, admission type) (Table 2, Table 3). Figure S2(A) shows eICU AUROC by age subgroup, and Figure S2(B) eICU AUROC by gender subgroups. A silent-trial analysis estimated lead time among alerted events. Lead time was defined a priori as the interval from the first model alert at a given horizon to the recorded time of in-hospital death for true-positive cases; for survivors, lead time is undefined and not analyzed. We included explainability summaries (global SHAP and case-level waterfalls) for transparency only (not used for decisioning) [7]. Internal data were drawn from MIMIC-IV; external validation used eICU-CRD [1, 8].

**Table 2.** External performance by window (eICU)

| Window (h) | N | Prevalence (%) | AUROC (95% CI) | AUPRC (95% CI) | Brier (95% CI) | ECE (95% CI) |
| --- | --- | --- | --- | --- | --- | --- |
| 6 | 19,182 | 12.5 | 0.857 [0.847, 0.866] | 0.546 [0.528, 0.565] | 0.077 [0.075, 0.080] | 0.046 [0.042, 0.049] |
| 12 | 18,523 | 12.2 | 0.864 [0.855, 0.873] | 0.535 [0.515, 0.555] | 0.076 [0.074, 0.079] | 0.023 [0.022, 0.026] |
| 18 | 17,245 | 12.4 | 0.835 [0.826, 0.844] | 0.453 [0.435, 0.472] | 0.087 [0.084, 0.089] | 0.044 [0.041, 0.048] |
| 24 | 15,274 | 13.0 | 0.825 [0.816, 0.834] | 0.432 [0.410, 0.450] | 0.091 [0.089, 0.094] | 0.036 [0.033, 0.041] |

*Contributing sites per window (k): 6 h = 36; 12 h = 33; 18 h = 32; 24 h = 29. Window cohorts were filtered independently to avoid horizon leakage.*
*AUROC/AUPRC (95% CI); Brier/ECE (lower is better); $\tau_{ij}$ = prespecified validation threshold; $\tau_{CGT}$ = guardrail threshold (alerts/100 admissions). **h** = hours.*

**Table 3.**
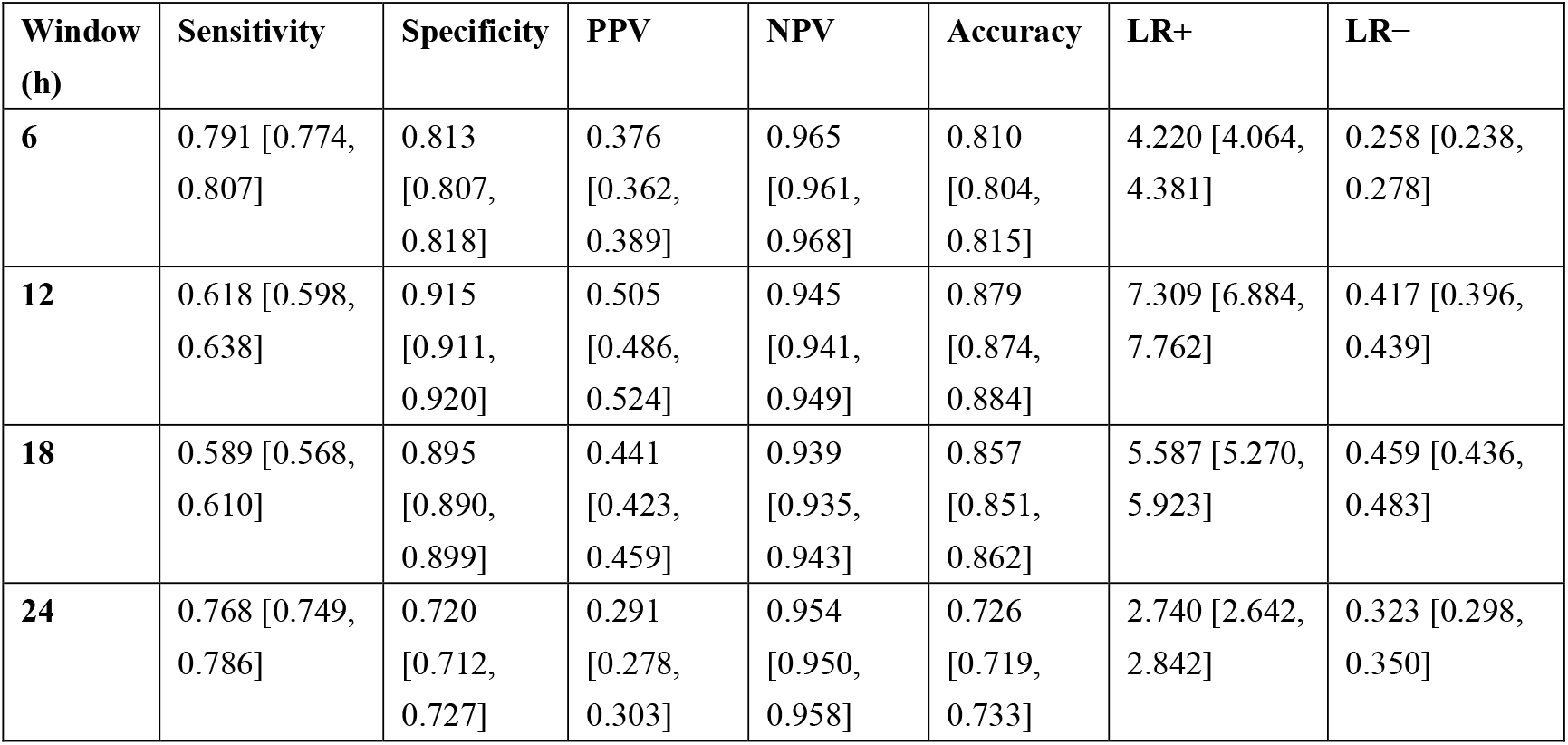
Prespecified operating-point metrics (95% CI)

| Window (h) | Sensitivity | Specificity | PPV | NPV | Accuracy | LR+ | LR– |
| --- | --- | --- | --- | --- | --- | --- | --- |
| **6** | 0.791 [0.774, 0.807] | 0.813 [0.807, 0.818] | 0.376 [0.362, 0.389] | 0.965 [0.961, 0.968] | 0.810 [0.804, 0.815] | 4.220 [4.064, 4.381] | 0.258 [0.238, 0.278] |
| **12** | 0.618 [0.598, 0.638] | 0.915 [0.911, 0.920] | 0.505 [0.486, 0.524] | 0.945 [0.941, 0.949] | 0.879 [0.874, 0.884] | 7.309 [6.884, 7.762] | 0.417 [0.396, 0.439] |
| **18** | 0.589 [0.568, 0.610] | 0.895 [0.890, 0.899] | 0.441 [0.423, 0.459] | 0.939 [0.935, 0.943] | 0.857 [0.851, 0.862] | 5.587 [5.270, 5.923] | 0.459 [0.436, 0.483] |
| **24** | 0.768 [0.749, 0.786] | 0.720 [0.712, 0.727] | 0.291 [0.278, 0.303] | 0.954 [0.950, 0.958] | 0.726 [0.719, 0.733] | 2.740 [2.642, 2.842] | 0.323 [0.298, 0.350] |

We conducted a retrospective observational study using de-identified electronic health records from two publicly available critical care databases. Development/internal validation: MIMIC-IV v2.2 (2008–2019) [9, 10]; external validation: eICU-CRD (2014–2015) [11, 12]. We adhered to each database’s usage guidelines and derived index ICU admissions for adults (≥18 y) with known in-hospital mortality; horizon-specific eligibility (details in paragraph S2). MIMIC-IV contains ICU admissions from a single academic medical center across multiple ICUs with rich, high-frequency clinical measurements and outcomes. eICU-CRD spans 208 ICUs across the United States with diverse settings and practice patterns, enabling assessment of transportability across sites. Cohort construction, feature extraction, and windowed prediction logic were harmonized between development and validation to preserve comparability across 6, 12, 18, and 24 h horizons after ICU admission. Feature importance across all windows is shown in Figure S4. We harmonized units/time-stamps across MIMIC-IV/eICU; full specifications in paragraph S1.

### Outcomes and Features

The binary endpoint was in-hospital mortality. For external validation, outcomes were taken as recorded in eICU-CRD discharge disposition and verified against hospital mortality flags when available. Horizon-specific labels were defined in the standard way: the target is the probability of in-hospital death, conditional on information accrued up to the horizon (6, 12, 18, or 24 h post-ICU admission). Importantly, windows were prospective in construction, no data after the horizon were used to form predictors at that horizon. To reduce label ambiguity, we excluded admissions with missing discharge status or inconsistent mortality indicators. Dataset/source indicators were used only for diagnostics and were excluded from model inputs. Early-ICU vitals/labs and demographics; leakage-prone variables excluded as well as in-fold imputation and no cross-split leakage are detailed in paragraph **S3**.

### Model Development and Validation

Preprocessing (median imputation) and any class-imbalance strategy were fit on the training split only. For imbalance, we used either scale-pos-weight tuning or SMOTE on the training matrix (never across splits), with the chosen approach logged per horizon. Predictors were standardized only when needed for baseline comparators (e.g., logistic regression); tree-based models used raw scales. Details of imbalance handled by prespecified rule; and isotonic calibration internally and external calibration are reported in paragraph **S5**.

### Calibration

We applied isotonic regression calibration separately for each horizon on the internal validation split and held the calibrated models fixed for external testing. Calibration was assessed in external validation using Brier score, expected calibration error (ECE; equal-frequency bins), and reliability plots. Calibration-in-the-large (CITL) and calibration slope were estimated with bootstrap confidence intervals. No recalibration was performed on eICU; external calibration is reported descriptively. Prespecified threshold set internally and held fixed (paragraph **S6**). Temporal blocks and site pooling performed (details in paragraph **S7**).

### Baselines

Throughout, we enforced leakage guards: (i) exclusion of post-horizon information (e.g., length-of-stay derived fields), (ii) strict in-fold fitting of imputer/SMOTE, (iii) fixed random seeds for splits, and (iv) reporting on external data without any threshold recalibration. We audited feature matrices before training to confirm that forbidden columns (e.g., icu_los_hours) and dataset indicators (source) were absent from X, and we re-checked the released artifacts accordingly. Regularized LR baseline included and leakage audits performed, details in paragraph **S8**.

### Statistical analysis

Continuous variables are reported as mean ± SD or median [IQR], and categorical variables as counts (%). Group differences between development and external cohorts were summarized descriptively. Discrimination was measured by AUROC and AUPRC at each horizon, with 95% CIs via nonparametric patient-level bootstrap (10,000 resamples). Because event prevalence is modest, AUPRC is reported alongside AUROC to reflect class imbalance. We summarize horizon-wise performance with a macro-average across 6/12/18/24 h (unweighted).

At the defined operating thresholds, we report sensitivity, specificity, PPV, NPV, accuracy, and likelihood ratios with 95% CIs (bootstrap; exact intervals guarded when percentile bounds exceeded [0,1]). Site-level AUROCs were pooled using a random-effects (DerSimonian–Laird) model with inverse-variance weighting; standard errors were derived from reported CIs when available, otherwise approximated via the Hanley–McNeil variance. We report pooled AUROC, 95% CI, I^2^, and number of sites per horizon.

## RESULTS

### Patient characteristics

We evaluated a windowed ICU mortality model at four horizons, 6, 12, 18, and 24 h after ICU admission, in an external eICU-CRD cohort using window-specific inclusion to prevent horizon leakage. Window-level performance summaries are in Table 2. External eICU cohorts per horizon with ~12–13% prevalence (see Table S1). Because deployment hinges on workload and timing, we quantified operational impact at the guardrail threshold. At 12 h, the alert rate was 61.4 per 100 admissions (PPV 0.304), translating to 5.1-10.2 clinician-hours per 100 admissions under 5-10 min/alert assumptions, with a median lead time of 138.6 h (IQR 54.6– 286.3) among alerted events; the base-case NMB peaked near τ_*CGT*_(Figure 3D; Table S4).

### Primary Performance

Discrimination was high across all windows and tapered mildly with longer horizons. Window-level AUROC (AUPRC) was 0.857 (0.546) at 6 h, 0.864 (0.535) at 12 h, 0.835 (0.453) at 18 h, and 0.825 (0.432) at 24 h (Figure 2A). Precision–recall curves remained meaningfully above the no-skill baseline (class prevalence ≈12–13%) (Figure 2C).

**Figure 1a.**
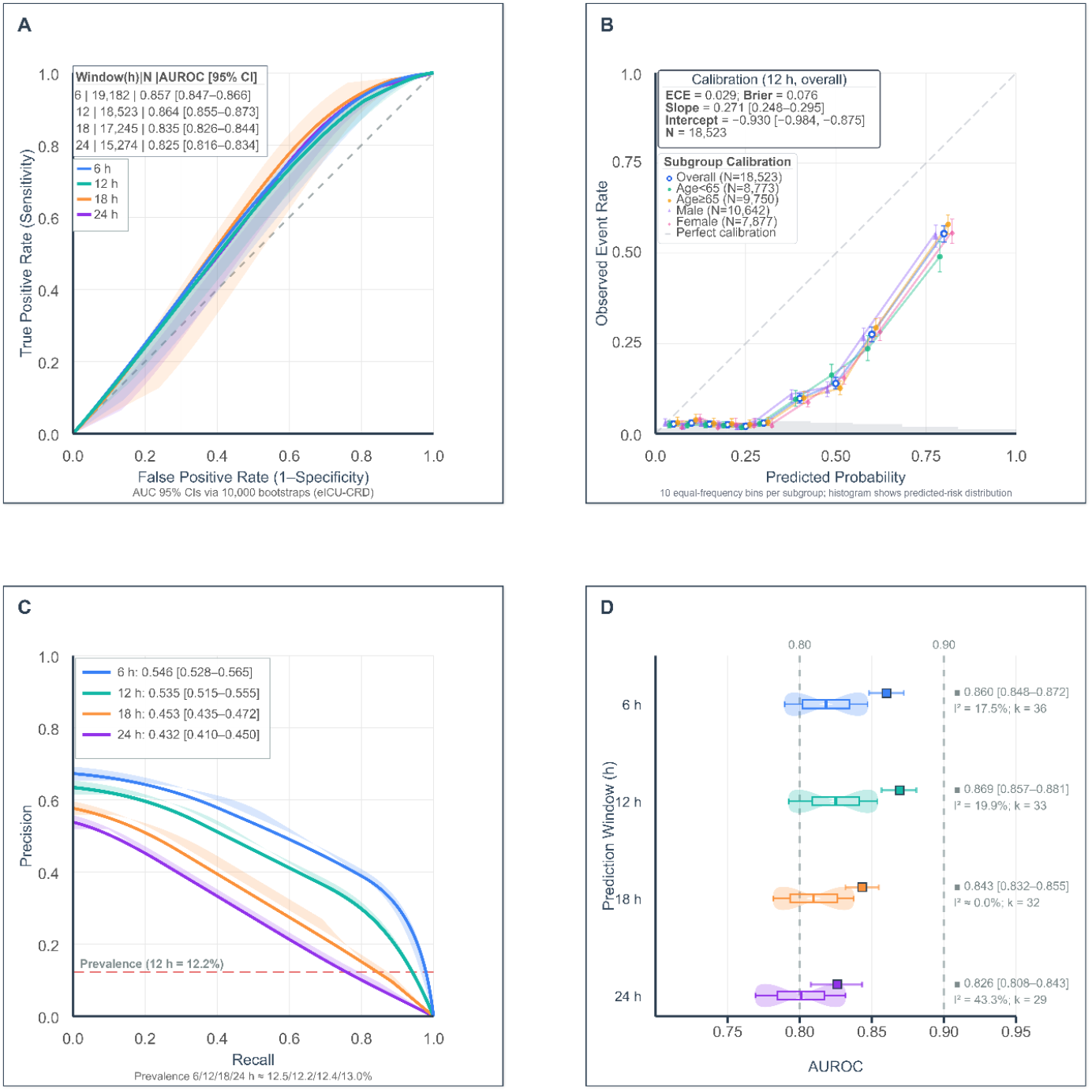
Model performance in external validation (eICU-CRD). **(A)** ROC curves by prediction horizon with 95% CIs. **(B)** Calibration plot at 12h showing observed vs predicted risk. **(C)** Precision-recall curves; dashed lines indicate outcome prevalence. **(D)** Site-level AUROC distributions with pooled estimates.

**Figure 2.**
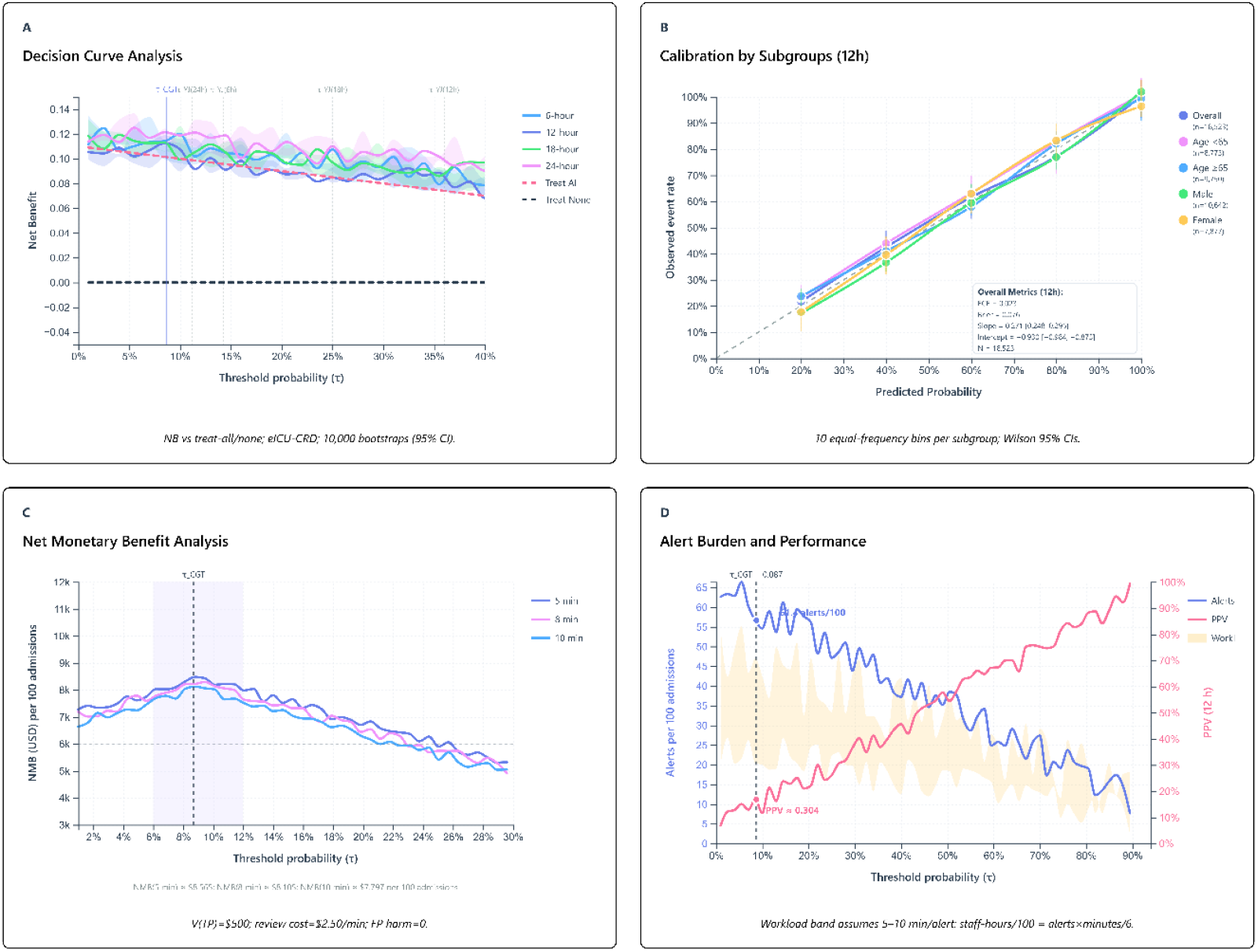
Clinical utility and operational metrics. **(A)** Decision curves showing net benefit vs treat-all/none strategies. **(B)** Subgroup calibration at 12h. **(C)** Net monetary benefit curves under different review time assumptions. **(D)** Relationship between alert threshold, workload, and PPV; vertical line marks operational guardrail (τ_*CGT*_=0.087).

### Calibration and Clinical Utility

Calibration and reliability were acceptable across horizons; at 12 h, CITL and slope are in Table S2, the calibration plot is in Figure 2B, horizon-wise reliability curves (6/12/18/24 h) are in Figure S1, and subgroup reliability (10 equal-frequency bins) is in Figure 3B. Decision curves (Figure 3A) showed positive net benefit across clinically relevant thresholds (0.01–0.30); at 12 h, net benefit exceeded treat-all/none in the mid-risk region and remained robust in sensitivity analyses [4, 7].

**Figure 3.**
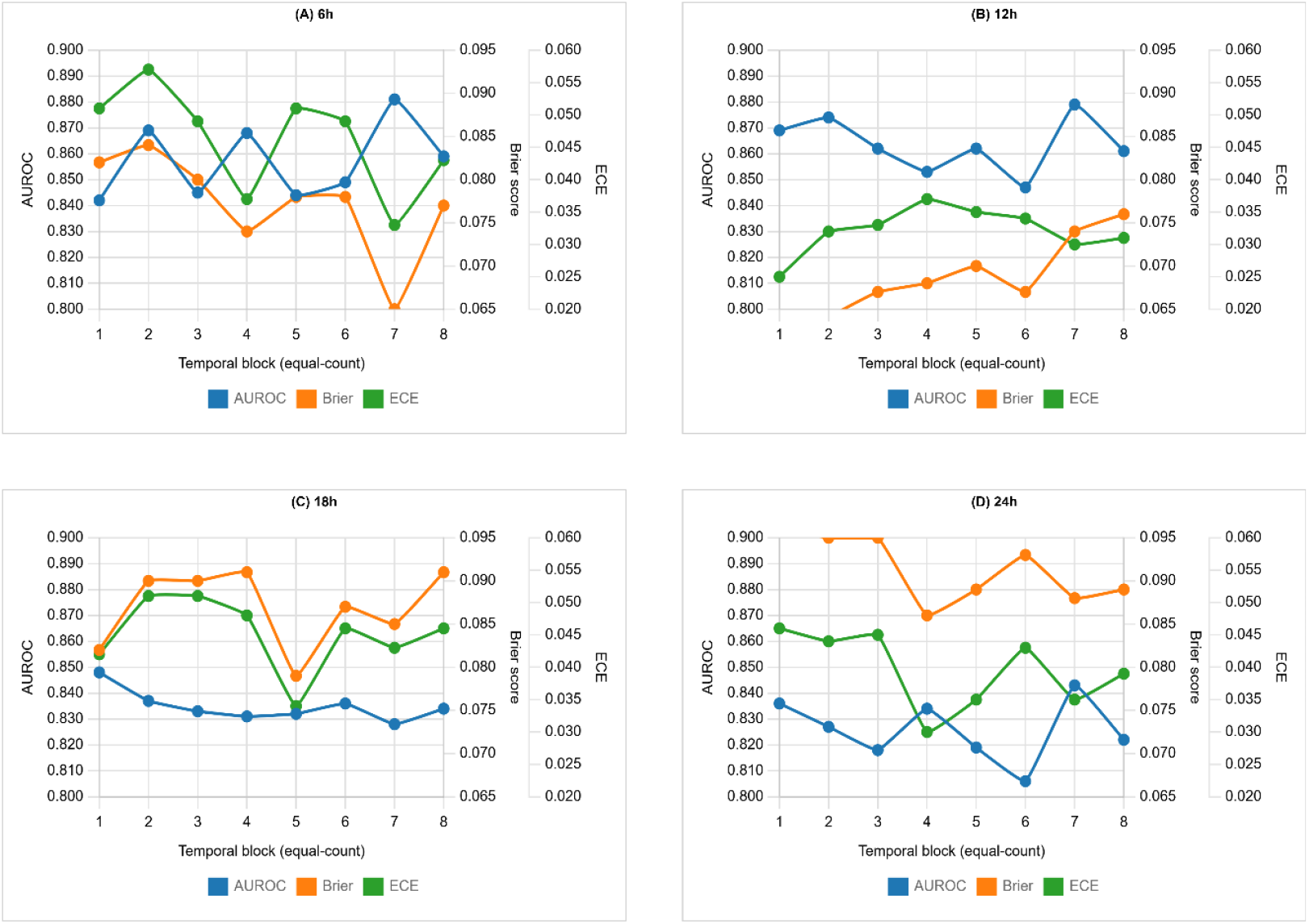
Temporal stability analysis. Performance metrics (AUROC, Brier score, ECE) across eight equal-count temporal blocks for each prediction horizon, showing stable discrimination without calendar-time degradation.

### Operating points and Subgroup stability (12 h)

The validation threshold τ_γj_ (chosen on calibrated risks in the internal split) was prespecified and carried unchanged to eICU-CRD. For operations, a 12 h guardrail τ_CGT_ targeted a predefined alert budget. In external testing, τ_γj_ was 0.143 (6 h), 0.360 (12 h), 0.250 (18 h), 0.112 (24 h). At 12 h, τ_γj_ yielded sensitivity 0.618, spec 0.915 (PPV 0.505; NPV 0.945; Table S3). At 12 h, threshold-fixed sensitivity/PPV showed modest age-related variation consistent with prevalence; full 95% CIs are in Table S5.

### Temporal Stability

Eight equal-count calendar blocks per horizon showed stable AUROC, Brier, and ECE without monotonic degradation (Figure 4A–D; Table S7; Figure S3).

### Site-level heterogeneity and Pooled discrimination

Random-effects meta-analysis showed consistently strong pooled discrimination across windows (AUROC ≈ 0.83-0.87), with low-moderate heterogeneity at 6-12 h, negligible heterogeneity at 18 h, and higher dispersion at 24 h. The 12 h window had the highest pooled AUROC (0.869, 95% CI 0.857-0.881), whereas the near-zero *I*^2^ at 18 h is compatible with within-site variance alone; the higher *I*^2^ at 24 h likely reflects later-stay missingness and practice variation. Full window-specific estimates (pooled AUROC with 95% CI, *k*, and *I*^2^) are shown in Figure 2D and detailed with site-level forest plots in Table S6. Random-effects pooling (DerSimonian–Laird) follows standard methods as described [13, 14].

### Operational impact at the Guardrail

At the 12 h guardrail (τ_CGT_ = 0.087), the model fired at 61.4 alerts/100 admissions with PPV = 0.304, yielding ~42 true and 96 false alerts per 1,000 admissions (Table S3; Table S4). In silent-trial assessment, alerts preceded event recognition by median 138.6 h (IQR 54.6–286.3), indicating substantial lead time before activation. Assuming 5-10 min review per alert, the review burden is ~5.1–10.2 clinician-hours/100 admissions. Under these assumptions, net monetary benefit (NMB) peaked near τ_CGT_. This explicit conversion from alert rate to staff time leading to economic proxy aligns with recommendations to verify burden and value during pre-deployment “shadow mode”/silent-trials [15, 16].

### Subgroup analyses

At τ_CGT_ and τ_*YJ*_, subgroup performance showed no clinically concerning drop-offs across sex, age bands, or admission type. Given operational salience, we also inspected a payer-type slice (Medicare/Medicaid/Private/Self-pay/Other) as an audit rather than a subgroup-optimized model; no material imbalances in precision or false-negative rates were observed at the guardrail point (Table S8). This aligns with calls to surface fairness-relevant outcomes during evaluation and governance of ICU AI [17].

### Model Interpretation

We include global SHAP (Figure S5) and exemplar case waterfalls (Figure S6) for transparency; explanations were not used for alerts or decisions [18, 19]. SHAP summaries (Figure S7) provide additional insight into how the model prioritizes variables across different time windows and SHAP dependency plots (Figure S8) illustrate how individual features interact and influence the model’s prediction at different levels.

## Discussion

We reported **equity audits** and **transparency** outputs; explanations were **not** used for decisioning [20, 21]. Post-deployment, we will monitor prevalence, risk distributions, AUROC/AUPRC, and calibration (slope/intercept, ECE) on rolling windows. Recalibration will be triggered if AUROC drops by ≥0.03 or ECE increases by ≥0.05 compared with the external-validation baseline. Our subgroup analyses suggest **stable sensitivity** across sex and clinically interpretable variation across age bands and admission sources at a **fixed, prespecified threshold**. PPV differences tracked subgroup prevalence rather than model collapse.

Across 6, 12, 18, and 24 h windows, discrimination sat within a narrow, clinically strong band, calibration was consistently tight by Brier and ECE, and decision-curve analysis showed positive net benefit across clinically plausible thresholds [21] **(Table 3)**. Operating-point behavior aligned with clinical intuition: earlier windows favored sensitivity and NPV, whereas the 12 h window favored specificity and PPV. For example, at 6 h (τ_γj_ = 0.143) sensitivity was0.79 with NPV 0.965; full, window-specific estimates with uncertainty are presented in Results. Together, these findings indicate stable early discrimination with strong rule-out performance and a predictable precision–recall trade-off as horizons lengthen. Using site-stratified meta-analysis on a large multicenter cohort, we observed low-to-moderate heterogeneity at 6 h and 12 h, minimal heterogeneity at 18 h, and moderate heterogeneity at 24 h; even at 24 h, pooled discrimination remained clinically strong **(Table S6)**. These data support cautious adoption with prospective monitoring and routine recalibration [13].

Temporal behavior was stable. Window-specific panels of AUROC, Brier, and ECE across equal-count temporal blocks showed no monotonic degradation; fluctuations were modest and centered on window means, suggesting that case-mix shifts during the evaluation period did not materially distort performance. The mild rise in Brier with longer horizons is consistent with greater outcome uncertainty as more future events are anticipated, while low ECE values indicate acceptable probability calibration in external validation. Taken together, small Brier and low ECE support using the probabilities without immediate recalibration in similar settings; however, we recommend ongoing monitoring and simple intercept/slope updates as needed.

Net-benefit gains clustered in the 5–20% risk band typical of early-warning and escalation workflows. At 6 h, the threshold reflects a conservative, sensitivity-forward stance appropriate when the downside of a missed deterioration is high and interventions are comparatively low risk. By 12 h, a higher threshold increases PPV and specificity, which is desirable when downstream actions (e.g., family meetings, imaging, transfers) have higher opportunity costs. The 18 h and 24 h operating points preserved clinically meaningful detection while tempering avoidable alarms as clinical context evolves. Importantly, bounding alerts per 100 admissions via a guardrail directly addresses the operational burden by aligning notifications with staffing capacity; the silent-trial analysis further supported feasibility, with alerts preceding event recognition by a substantial median lead time. Consistent with Methods, the net monetary benefit (NMB) scenario peaked near the guardrail under the 5-10 min/alert handling assumption; assumptions and sensitivity ranges are detailed in Methods, and outcomes are summarized in Results.

Our analysis hews to contemporary reporting norms, emphasizing calibrated risk and threshold-level trade-offs over rank discrimination alone, and adds deployment-oriented elements that clarify what it would cost to run the service and how much signal it would return [14]. Beyond AUROC, we report Brier/ECE, operating-point metrics with uncertainty, and decision curves; we enforce leakage guards (exclusion of length-of-stay proxies, strict in-fold preprocessing, per-horizon calibration). Unlike single-time-point score validations, the fixed-horizon lens clarifies when the model is most actionable and how thresholds should be tuned to workload and harm preferences at different times in the stay.

This study has limitations. It is a retrospective validation on de-identified multicenter data; residual confounding, undocumented workflow differences, and selection effects remain possible. The window definitions are pragmatic and do not capture continuously updated predictions; they should be viewed as deployable checkpoints aligned with admission-to-early-stay workflows. The feature set is constrained to routinely available variables; unmeasured confounders (e.g., clinician gestalt, imaging) may limit ceiling performance. While calibration transferred well, local intercept/slope updates at go-live are prudent, especially where baseline mortality or admission practices differ. Finally, moderate heterogeneity at 24 h warrants local monitoring and, if necessary, site-specific threshold tuning.

Clinical implications follow directly. For early screening (6–12 h), sensitivity-oriented thresholds that maximize NPV and net benefit pair well with low-harm, reversible care pathways (enhanced monitoring, early review). For later checkpoints (18–24 h), resource-aware thresholds can reduce avoidable alarms while preserving detection, particularly when capacity is tight. Prospective deployment should include drift monitoring (risk distribution, prevalence, calibration slope/intercept), pre-registered operating points, and lightweight recalibration triggers [13]. Future work will extend beyond fixed horizons to rolling predictions, add fairness and subgroup stability analyses, and test prospective impact on workflow and outcomes.

## Conclusions

From 6-24 h after ICU admission, the model showed strong discrimination, small calibration error, low-to-moderate site heterogeneity, and consistent decision-curve benefit. Framing outputs as a staffable alert program, via guardrails, workload translation, silent-trial lead time, and a succinct economic view, clarifies what it would cost to run the service and how much signal it returns at a chosen threshold. These properties support careful clinical piloting with routine monitoring and recalibration.

## Data Availability

Source code for the Sentinel-ICU analyses and demo services is available at GitHub (https://github.com/Sjtu-Fuxilab/Sentinel-API). An archived snapshot of the exact version used here, including model artifacts, will be deposited on Zenodo and cited with a DOI upon acceptance. Two runnable packages accompany this submission: Supplementary Tool S1, the Sentinel-ICU Mortality Inference API (FastAPI, v1.0), and Supplementary Tool S2, the net monetary benefit (NMB) forecaster app (FastAPI, v2.3). Mirror copies of both tools are hosted in the same GitHub repository.

## Abbreviations

CI: Confidence interval
CITL: Calibration-in-the-large
DCA: Decision-curve analysis
ED: Emergency department
ECE: Expected calibration error
HER: Electronic health record
FP / FN: False positive / false negative
ICU: Intensive care unit
IQR: Interquartile range
K: Number of sites included in a pooled/meta-analysis at a horizon
LOS: Length of stay
NMB: Net monetary benefit
NPV / PPV: Negative / positive predictive value
PR: curve Precision–recall curve
ROC: Receiver-operating characteristic
SD: Standard deviation
SE / SP: Sensitivity / specificity
τ_γj_: Youden’s J on calibrated risks, fixed from internal validation
τ_CGT_: Clinical guardrail threshold
TP / TN: True positive / true negative
V, C, W, h: Parameters in NMB: value per TP, cost per FP, staff cost/min, review minutes per alert

## Data Availability

Data are publicly available on PhysioNet under credentialed access. We used MIMIC-IV v2.2 (DOI: 10.13026/6mm1-ek67) and eICU-CRD v2.0 (DOI: 10.13026/C2WM1R). Please also see the dataset descriptor articles in *Scientific Data* for MIMIC-IV (2023) and eICU-CRD (2018). PhysioNet platform citation: Goldberger et al., *Circulation* 2000 [22].

## Code Availability

Source code for the Sentinel-ICU analyses and demo services will be available at GitHub (https://github.com/Sjtu-Fuxilab/Sentinel-API). An archived snapshot of the exact version used here, including model artifacts, will be deposited on Zenodo and cited with a DOI upon acceptance. Two runnable packages accompany this submission: **Supplementary Tool S1**, the Sentinel-ICU Mortality Inference API (FastAPI, v1.0), and **Supplementary Tool S2**, the net monetary benefit (NMB) forecaster app (FastAPI, v2.3). Mirror copies of both tools are hosted in the same GitHub repository.

## Ethics approval and consent to participate

This study used de-identified public datasets (MIMIC-IV v2.2 and eICU-CRD). According to our institutional policy, the analysis constitutes non-human subjects research and was exempt from IRB review. No patient consent was required.

## Authors’ Contributions

**SAZ** conceived and designed the study; acquired and curated the data; implemented the software; performed the analyses; created the figures/tables; and drafted the manuscript. **WQ** supervised the study, contributed to study design and methodological oversight, provided resources, critically revised the manuscript, and served as the **corresponding author**; **WQ** also secured **funding. TAS** reviewed the manuscript and contributed to design and methodology. **AAK** provided clinical interpretation and validation and critically revised the manuscript. **MSF** provided clinical oversight and resources, and critically revised the manuscript. **All authors read and approved the final manuscript**.

## Consent for publication

Not applicable.

## Clinical trial number

Not applicable.

## Supplementary Information

Supplementary Tables S1-S8 and Figures S1-S8 are available online.

